# Age-specific income losses due to HPV-attributable cancers in Singapore

**DOI:** 10.64898/2026.04.16.26351014

**Authors:** Robin Blythe, Nicholas Graves, N. Gopalakrishna Iyer, Marco A Peres

## Abstract

**Introduction:** The link between Human Papillomavirus (HPV) and cancer is well-established. In Singapore, bivalent HPV vaccines are subsidised for females, but not males. Economic analysis of HPV vaccination has generally assessed the costs to the health system, but this may not be as relevant to individual decision-making as potential lost income. We estimated the impact of bivalent HPV 16/18 vaccination on sick leave, unemployment, and premature mortality as a function of age and sex to understand the broader impact of HPV-related cancers.

**Methods:** We developed a population-level economic model to estimate lifetime income losses by diagnosis age, sex and cancer type. We applied sex- and cancer-specific Cox regressions to the Singapore Cancer Registry for annual predicted survival from 1992 to 2022. These were combined with census and employment data to estimate HPV-associated income losses in Singapore. Attributable fractions and vaccine effectiveness data for HPV 16/18 from the literature were used to estimate the effectiveness of bivalent HPV vaccination. Structural sensitivity analysis examined the role of 80% population coverage conferring herd immunity.

**Results:** The registry contained 17,294 individuals with an HPV-associated cancer diagnosis. Lost income was greatest for cervical cancer due to its high prevalence, however the losses per diagnosis were highest for oropharyngeal cancer. Bivalent HPV vaccination led to income benefits of $SGD1,397 [$895 to $1,838] in girls and -$62 [- $76 to -$48] in boys. A gender-neutral HPV vaccination of 80% of 15-year-old Singaporeans, conferring herd immunity, would have lifetime income protective benefits of $24.4m [$14.2m, $33.7m] per cohort, a five-fold return on investment.

**Conclusions:** In addition to avoiding healthcare costs and lost quality of life, parents should consider vaccination as a means of avoiding potential income losses. A national policy of gender-neutral HPV vaccination could deliver substantial income protection due to both individual vaccine protection and herd immunity.

## Introduction

Human Papillomavirus (HPV) is a common risk factor for cancers of the cervix, oropharynx, anus, and genitals. Infection with high-risk HPV variants can lead to lesions, which may progress to cancer.^1^ Cervical cancer is the most common, accounting for 80% of HPV-related cancers;^2^ globally, HPV-related cancers made up 4.4% of all new cancer cases in 2020.^3^ Cervical cancer was the fourth leading cancer in females globally in 2022, with over 660,000 (7%) of all new cases, and nearly 350,000 deaths.^4^ In addition to mortality, HPV-related cancers can lead to significant disability, including loss of fertility and low self-esteem in cervical cancer survivors^5^ and facial disfigurement,^6^ trouble speaking and trouble swallowing^7^ in oropharyngeal cancer survivors.

Vaccination for HPV can prevent infection with the virus and is consequently crucial for cancer prevention,^8^ in addition to using contraception and reducing the number of sexual partners.^9^ In Singapore, bivalent HPV vaccination (covering two carcinogenic HPV strains) in young women is encouraged to prevent cervical cancer. It is free of charge in schools and polyclinics for female citizens until age 17, and heavily subsidised from ages 18 to 26.^10^ Nonavalent (covering nine HPV strains, including those responsible for genital warts) vaccines are available at polyclinics and private practices, but not subsidised by the government, and the quadrivalent vaccine is no longer available as of 2025. Uptake in young girls has increased considerably over time, with a 2023 study estimating bivalent coverage of 93%, up from 14% in 2014, and just 2% of school-aged girls opt out of HPV vaccination.^11^ Bivalent and quadrivalent, but not nonavalent, vaccines have been deemed cost-effective in Singapore for the prevention of cervical cancer in females.^12–14^

In males, however, vaccination is neither endorsed nor subsidised, despite reducing risks of HPV transmission and rates of oropharyngeal and anogenital cancers.^15^ Vijayalakshmi & Goei (2022) suggest that this is primarily because vaccination is only considered for the prevention of cervical cancer.^11^ Wahab et al (2023) showed that the bivalent vaccine could be cost-effective in boys for the prevention of other HPV-related complications, depending on the discount rate applied to future costs.^16^ However, only the nonavalent vaccine is currently available for use in boys, which was highly unlikely to be cost-effective due to high vaccine costs.

While most cost-effectiveness analyses focus on the prevention of healthcare costs through vaccination, broader personal and societal costs are rarely addressed.

Marsh et al. reported that these costs, including the economic costs of long-term unemployment due to HPV-associated cancers, are often missing from economic evaluations.^17^ A more comprehensive understanding of these costs may also be important for increasing public knowledge of HPV vaccines, which are not well-understood in Singapore.^18^ Individuals are often insulated from healthcare costs by insurance or public financing, which may have an adverse influence on decision-making.^19^ Through understanding the personal costs of HPV-related cancers, individuals may be able to make more informed decisions about vaccination, and policymakers can use this evidence to design co-payments and public education in national vaccination programmes.

These costs may differ by the types of complications experienced by individuals with HPV, and will therefore likely differ by age between males and females.

Understanding these costs may be particularly informative for cancers that affect younger adults, including those associated with HPV. ^20^

In this economic modelling study, we examined the personal costs of HPV-associated cancers on lost income by age and sex in Singapore to assist in vaccine policy decision-making.

## Methods

### Study overview and data sources

We conducted a simulation study using age- and sex-specific risks of cancer diagnosis and mortality, employment, and median income. We obtained model parameters for cancer mortality from Singaporean cancer registry data. We combined cancer registry and census data to estimate the sex- and age-specific prevalence of each cancer in the Singaporean population. Employment data and median income were obtained from publicly available census data in 5-year age categories and transformed into age- and sex-specific weighted incomes using smoothing splines. In all cases, sex variables were extracted as reported in the different data sources described above.

We supplemented these data with estimates of sick leave following cancer diagnosis, vaccine efficacy and the proportion of cancer cases attributable to HPV from published literature. All data preparation, analysis, and figures were developed in R version 4·2·2.^21^ The model structure is shown in Figure 1. Model input parameters are described in Table 1; all data and code can be found at https://github.com/robinblythe/HPV_CEA.

**Figure 1:**
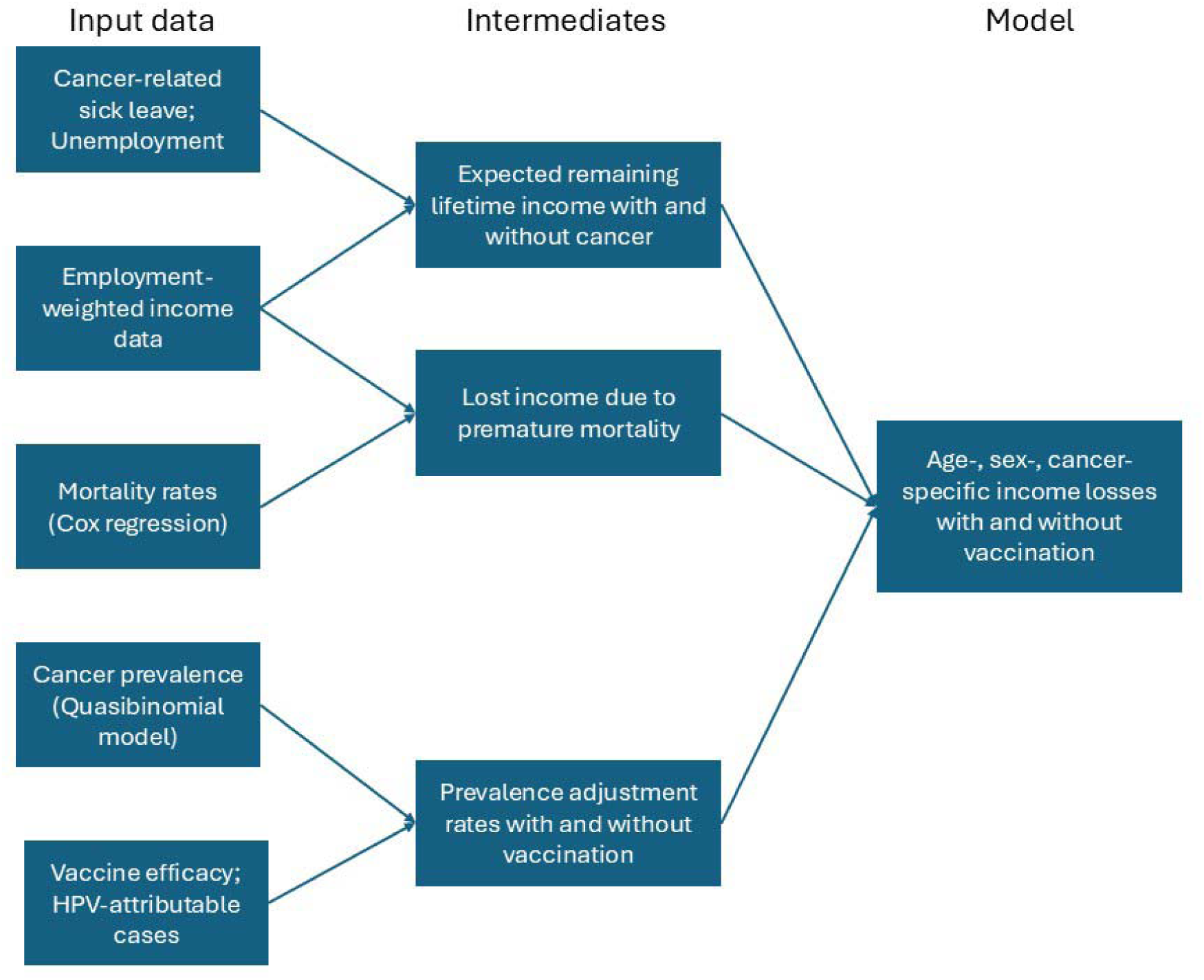
Simulation model structure. Intermediates refer to values used by the model to estimate total income losses that are also reported in the results as standalone quantities. HPV: Human Papillomavirus.

**Table 1:** Simulation model input parameters. Each parameter was sampled 10,000 times for the main analysis. The herd immunity parameter was sampled 1,000 times for the structural sensitivity analysis.

| Parameter | Point estimate | Distribution function | Source |
| --- | --- | --- | --- |
| Odds ratio of being employed after cancer diagnosis |  |  | de Boer et al (2009)<sup>27</sup> |
| Oropharyngeal | 0.27 | $\frac{\frac{Beta(75, 62)}{1 - Beta(75, 62)}}{\frac{Beta(116, 26)}{1 - Beta(116, 26)}}$ | Nasopharyngeal (proxy) |
| Female reproductive | 0.64 | $\frac{\frac{Beta(671, 648)}{1 - Beta(671, 648)}}{\frac{Beta(816, 506)}{1 - Beta(816, 506)}}$ | Female reproductive |
| Male reproductive | 0.57 | $\frac{\frac{Beta(423, 275)}{1 - Beta(423, 275)}}{\frac{Beta(1770, 659)}{1 - Beta(1770, 659)}}$ | Prostate (proxy) |
| Unpaid sick leave duration due to cancer diagnosis (months) |  |  |  |
| Oropharyngeal* | 2.75 | $\frac{\Gamma(3.545, 3.972) - 2}{4.345}$ | Baxi et al (2016)<sup>30</sup> |
| Female reproductive** | 6.96 | $\frac{\Gamma(148.905, 1.088) - 14}{30.438}$ | Roelen et al (2011)<sup>29</sup> |
| Male reproductive** | 3.53 | $\frac{\Gamma(659.678, 0.159) - 14}{30.438}$ | |
| Attributable fraction to HPV 16/18 |  |  |  |
| Cervical | 0.70 | $Beta(37000, 16000)$ | de Martel et al (2020)<sup>2</sup> |
| Anal | 0.75 | $Beta(30000, 10000)$ | |
| Vaginal | 0.49 | $Beta(7400, 7600)$ | |
| Vulval | 0.18 | $Beta(6200, 27800)$ | |
| Penile | 0.35 | $Beta(9100, 16900)$ | |
| Oropharyngeal | 0.26 | $Beta(24621, 71379)$ | |
| Probability of invasive cancer after vaccination |  |  |  |
| Cervical | 0.09 | $Beta(0.773, 7.787)$ | Lei et al (2020)<sup>33</sup> |
| Oropharyngeal | 0.11 | $Beta(0.981, 7.770)$ | Herero et al (2013)<sup>34</sup> |
| Female reproductive | 0.05 | $Beta(0.930, 18.548)$ | De Sanjose et al (2014)<sup>35</sup> |
| Male reproductive | 0.16 | $Beta(7.524, 39.539)$ | |
| Relative risk of infection due to herd immunity | 0.30 | $1 - Beta(2.975, 1.275)$ | Brisson et al (2016)<sup>36</sup> |
| Discount rate | 0.03 | Fixed | Singapore Ministry of Health<sup>28</sup> |

*\*Distribution measured in weeks*
*\*\*Distribution measured in days*

This study has received a “review not required” determination from the National University of Singapore Institutional Review Board (NUS-IRB-2025-265). Patient data were last accessed on 17 March 2025. While data within the registry were identifiable, analysis outputs were checked by the Singapore National Registry of Diseases Office to confirm non-identifiability prior to being extracted from the registry.

### Mortality estimates

We first developed a Cox regression to estimate mortality using data from the Singaporean Cancer Registry.^22^ We included all cancer diagnoses in Singapore from first use of the International Classification of Diseases for Oncology (ICD-O) for histology coding in 1992 until 2022, the latest update of the registry, to ensure a consistent study population. Cancer types were identified by their ICD-O label site codes (2^nd^ and 3^rd^ edition) corresponding to cancers of the oropharynx, anus/anal canal, vulva, vagina, cervix, and penis. The outcome variable was death due to cancer of the primary site.

Survival time was calculated as the time from diagnosis to death, censoring patients who were still alive as of 2022 or died from other causes. A restricted cubic spline with 4 knots was used to model the non-linear effect of age on mortality, and sex was included as a binary variable. The model was stratified based on cancer type to account for potential differences in the baseline hazard of each diagnosis. The regression was developed using the `survival` package in R.^23^

The regression equation was used to predict survival probabilities with standard errors up to 30 years from diagnosis for each age, cancer type, and sex. Estimates and standard errors were then converted to a set of Beta distribution parameters using the method of moments.

### Annual cancer prevalence estimates

To obtain a probability of cancer diagnosis for each age, sex, and cancer type, we obtained the average number of annual diagnoses at each age from the registry between 1992 and 2022, after confirming the trend was roughly stationary during this period. We then divided these mean values by the estimated population by age and sex in Singapore as of the latest census estimates (2024).^24^ To obtain smoothed estimates with standard errors, we fit a quasibinomial model for each cancer type and sex, with age at diagnosis as a predictor variable, and predicted the probability of diagnosis of each cancer at each age for both males and females.

### Income, employment and cancer-related labour force participation

Labour force participation and median incomes were obtained from the Singapore Ministry of Manpower and the Singapore Department of Statistics.^25,26^ As these statistics are published in 5-year bands by sex, we obtained annual estimates by age using smoothing splines, selecting a spar term based on visual inspection of the smoothed curves compared to the original data. We then multiplied the annual median income by the labour force participation at each age to obtain a weighted income value.

To account for unemployment due to each type of cancer, we obtained estimates from a meta-analysis published by de Boer and colleagues.^27^ The authors reported the number of individuals who were employed and unemployed for all cancers, including female reproductive cancers, prostate cancer and nasopharyngeal cancers. We applied unemployment data for nasopharyngeal cancer as a proxy for oropharyngeal cancer, the data for female reproductive cancer as the risk for anal, cervical, vaginal and vulval, and the data for prostate cancers as a proxy for the risk from male anal and penile cancers. We used the number employed and unemployed as alpha and beta parameters for Beta distributions, respectively, then converted these to odds ratios and multiplied them by the odds of employment for the population to get cancer-adjusted participation. This was multiplied by income to obtain a weighted income value for each type of cancer by age and sex.

As we aimed to estimate total income losses by age, we calculated the expected value of an individual’s remaining lifetime income for each age and sex. A 3% discount rate was applied to estimate the net present value of each year of income, as recommended by the Singapore Ministry of Health.^28^ Each discounted annual income value was added together to estimate the remaining participation-weighted income expected until age 84 (including retirement).

We also included estimates for the duration of sick leave accumulated during cancer treatment prior to affected adults returning to work. We obtained estimates from Roelen et al. (2011) reporting the number of days taken by patients diagnosed with female and male genital cancers,^29^ while estimates from Baxi et al (2016) were used for oropharyngeal cancer.^30^ These values were converted to months and used to parameterise Gamma distributions using the ShinyPrior tool.^31^ Duration of sick leave in months, less an assumed 14 days of statutory paid leave, was multiplied by the weighted monthly income for each type of cancer patient to obtain income losses due to sick leave.

### Vaccination

For the proportion of cases attributable to HPV 16/18, the strains targeted by the bivalent vaccine, we used estimates from de Martel et al.^32^ and converted these estimates to Beta distributions. We used bivalent vaccine efficacy values from three different data sources for cervical,^33^ oropharyngeal,^34^ and genital/anal cancers.^35^ The total cost of bivalent vaccination was taken from Phua et al. and updated to SGD 2025 (fixed at $138).^13^

### Model structure and probabilistic sensitivity analysis

To propagate sampling uncertainty throughout the model, we sampled from distributions of model input parameters 10,000 times, reporting the median and 95% uncertainty intervals. The model sampled each input distribution once per iteration to obtain different expected values of cancer diagnosis for each age of diagnosis from 16 to 84 years by sex and cancer type.

Sick leave and mortality were incorporated into the model as decrements to be subtracted from the total expected remaining lifetime income following cancer diagnosis. The difference between the lifetime expected income for a healthy individual and an individual diagnosed with cancer was reported in the results as the total income losses due to cancer at each age of diagnosis. This was then weighted by the probability of cancer diagnosis with and without vaccination. Vaccination benefits across cancer types were aggregated prior to subtracting the cost of vaccination as the treatment is preventive for all cancer-related complications of HPV infection.

### Model sensitivity to herd immunity

The model described above estimated the reduction in income losses from vaccination compared to no vaccination. However, any national HPV policy would likely lead to herd immunity with a sufficiently high number of vaccinated individuals. Over 1,000 simulations, we assessed structural model uncertainty and its effects on HPV policy by estimating the effect of herd immunity. A meta-analysis by Brisson et al (2016) assessed the effect of herd immunity and vaccine duration, finding that even if immunity lasted only 20 years, a 70% [95% CI: 21% to 100%] relative risk reduction in HPV would be anticipated over the lifetime of the vaccinated population given 80% coverage.^36^ This was incorporated into the analysis as a multiplier to the diagnosis probability, fit using a Beta distribution (Table 1). The overall population impact was assessed as costs and estimated income benefits at 80% vaccine coverage.

## Results

There were 17,294 patients in the registry with a diagnosis of HPV-associated cancer. Of these patients, 2,523 (15%) had a cancer-related death recorded in the registry. The average number of cases per year for each type of cancer as a function of age were few, averaging around 0·2 cases per year each, apart from cervical cancer, which averaged around 6·7 cases per year (Figure 2).

**Figure 2:**
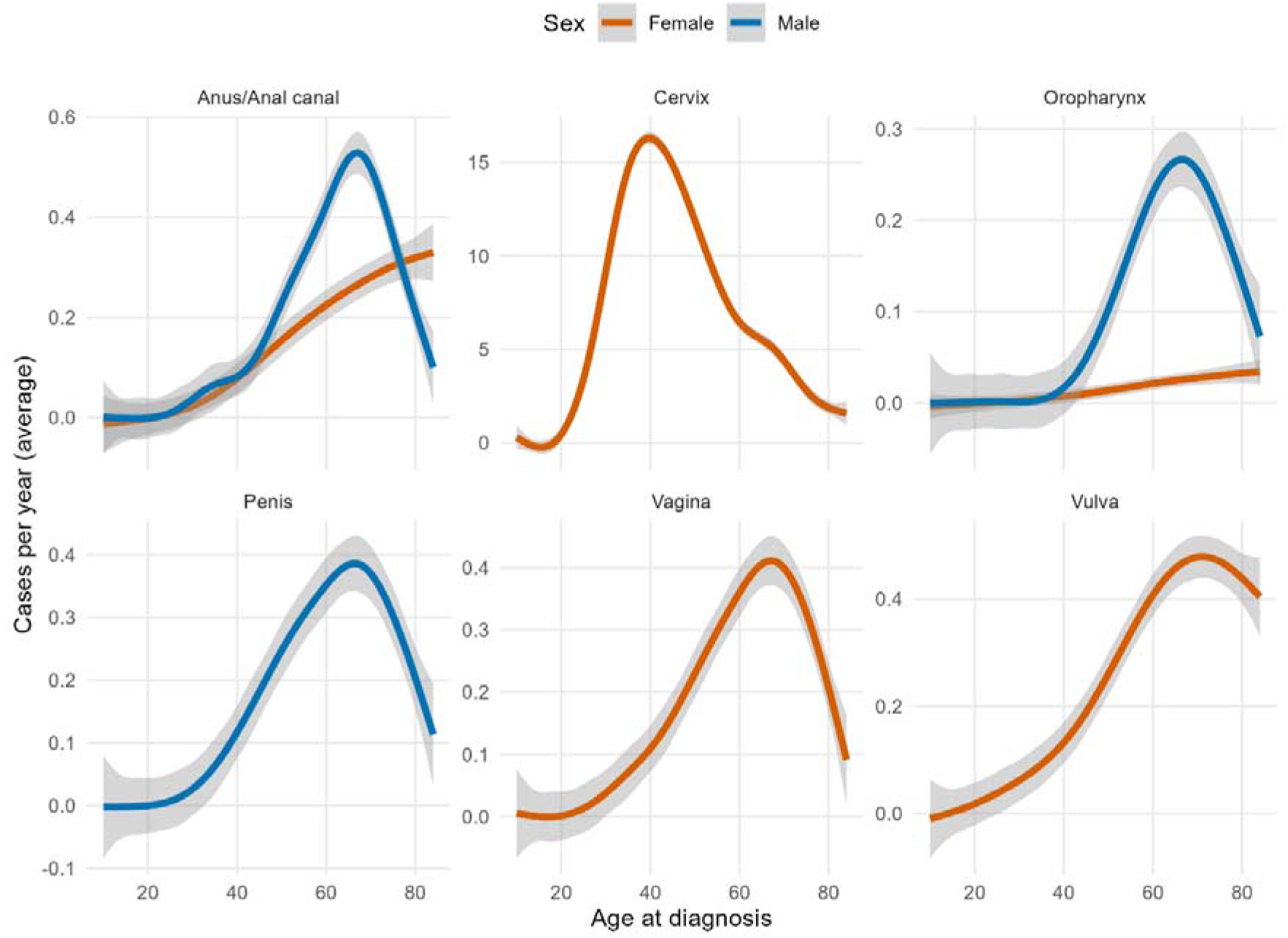
Average annual number of cases by sex and primary cancer site as a function of age.

Oropharyngeal cancer diagnosis led to the lowest survival of all HPV-associated cancers after adjustment for age and sex. However, it also represented the smallest share of HPV-associated cancers in Singapore (1·5%), and had the second-lowest attributable fraction to HPV 16/18 after vulval cancer. By contrast, cervical cancer, by far the most common cancer diagnosis representing 86% of the dataset, had the highest survival and youngest age at diagnosis. Adjusted survival curves for the Cox regression are shown in Figure 3.

**Figure 3:**
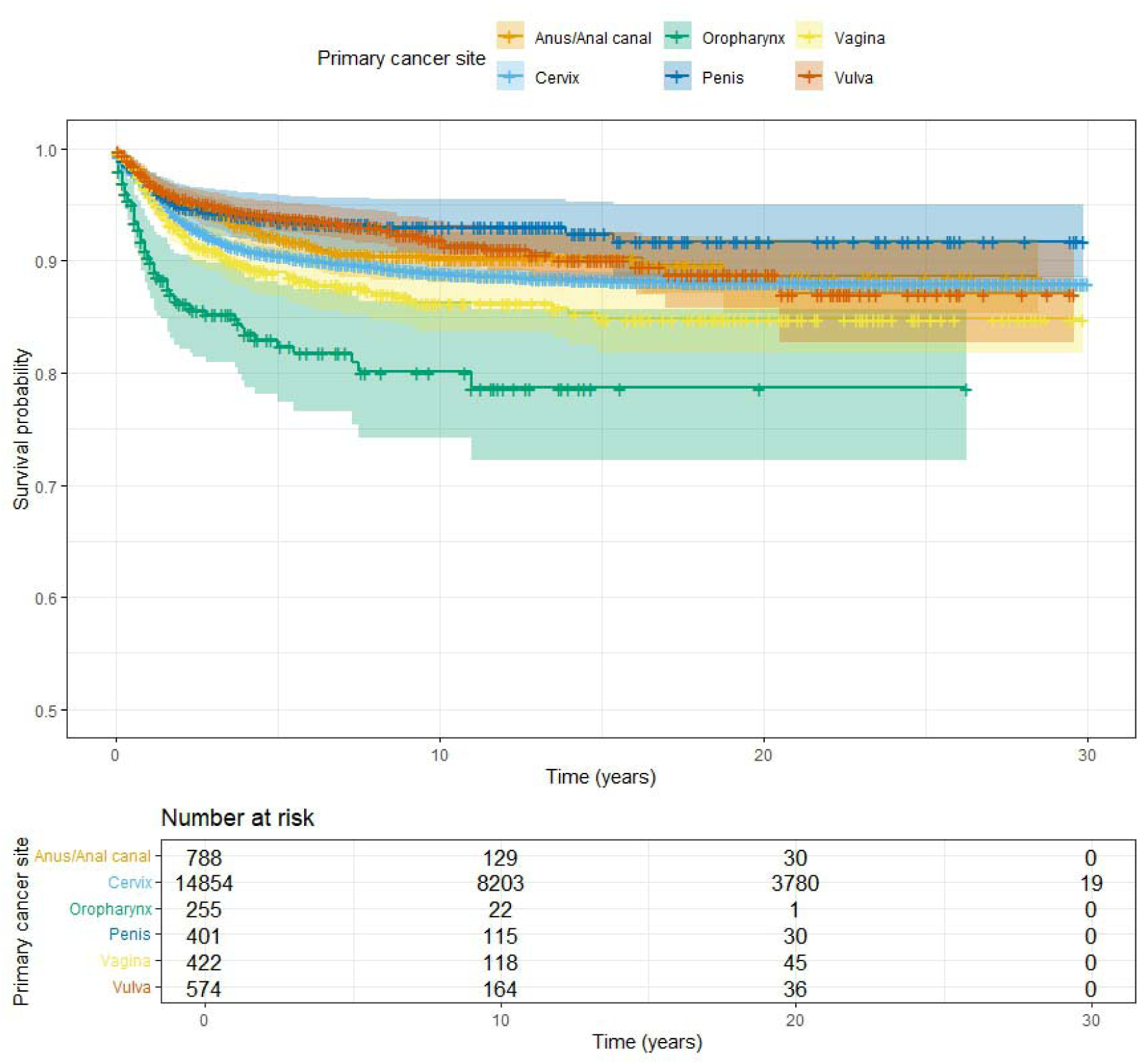
Adjusted survival curves from the Cox regression over 30 years. Each tick on the survival curve denotes a censoring event. For example, if patients were diagnosed in 2015 and followed until the last year of the dataset (2022), there would be a tick mark at 6 years.

Income loss as a function of diagnosis age (Figure 4), increased with each year younger an individual was when diagnosed with an HPV-related cancer, though this relationship was non-linear. Lifetime income losses for a range of diagnosis ages are shown in Table 2. Across both males and females, oropharyngeal cancer diagnosis led to the greatest income losses.

**Figure 4:**
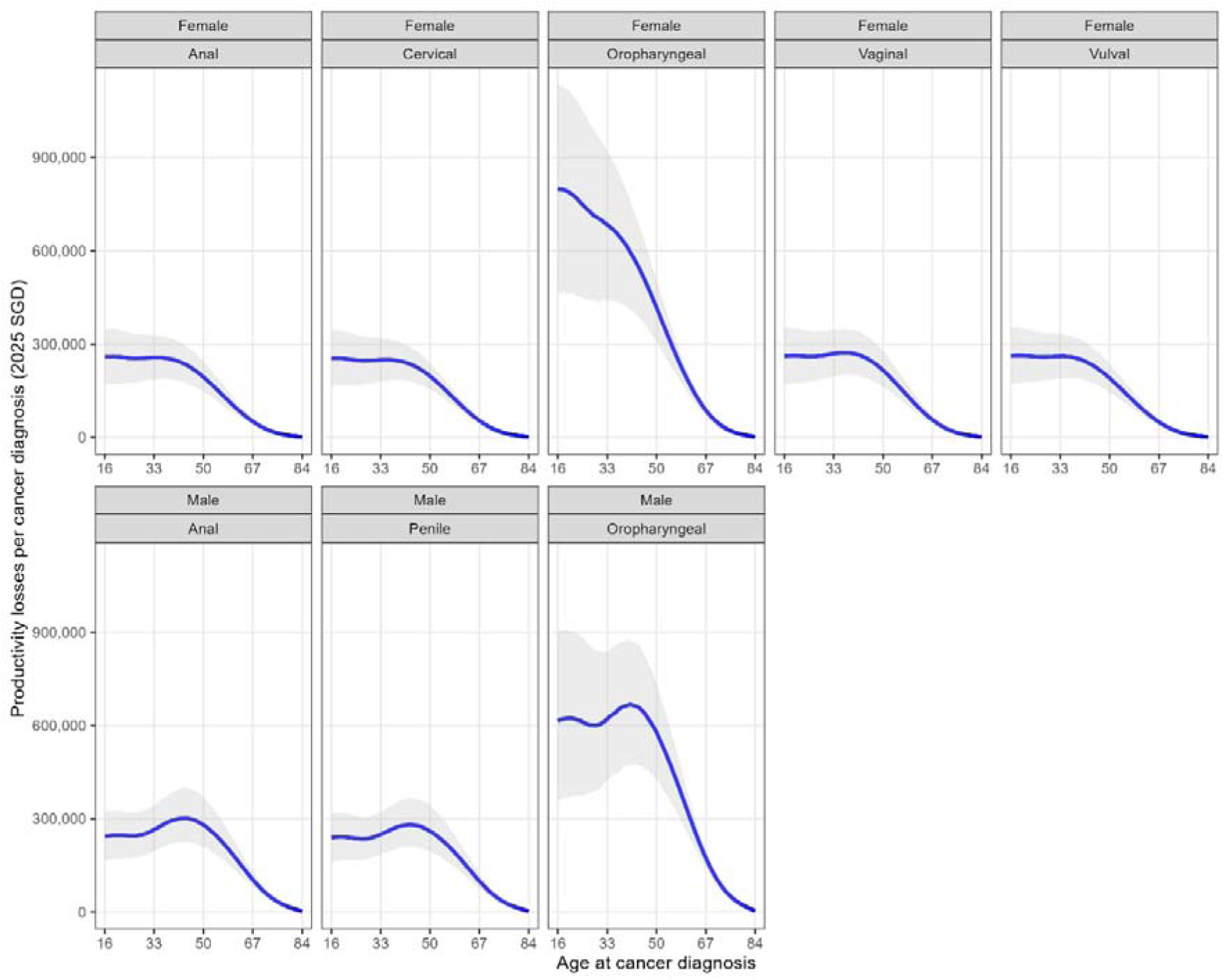
Predicted lifetime losses in income due to HPV-associated cancer diagnosis. The blue line reflects the median value, while the grey ribbon contains the 95% uncertainty interval.

**Table 2:** Median income losses in SGD by sex and primary cancer site for 3 selected ages at diagnosis: 33, 50 and 67. Lost income is calculated by diagnosis and assumes the diagnosed individual is earning the median income for their age. SGD: Singapore dollars.

| **Cancer diagnosis** | **Sex** | **Age** | **Median lost income** | **95% uncertainty interval** |
| --- | --- | --- | --- | --- |
| Anal | Female | 33 | \$256,403 | [\$185,668 to \$328,793] |
| | | 50 | \$193,629 | [\$147,847 to \$243,765] |
| | | 67 | \$50,953 | [\$39,768 to \$62,627] |
| | Male | 33 | \$264,897 | [\$196,273 to \$342,888] |
| | | 50 | \$280,106 | [\$210,983 to \$370,202] |
| | | 67 | \$104,550 | [\$81,827 to \$129,835] |
| Cervical | Female | 33 | \$249,017 | [\$179,780 to \$320,422] |
| | | 50 | \$196,720 | [\$154,766 to \$237,305] |
| | | 67 | \$53,677 | [\$44,486 to \$62,534] |
| Oropharyngeal | Female | 33 | \$683,556 | [\$438,361 to \$918,486] |
| | | 50 | \$417,139 | [\$307,611 to \$506,552] |
| | | 67 | \$87,280 | [\$70,140 to \$99,960] |
| | Male | 33 | \$620,485 | [\$421,639 to \$838,324] |
| | | 50 | \$578,634 | [\$425,213 to \$733,119] |
| | | 67 | \$174,160 | [\$140,784 to \$200,951] |
| Penile | Male | 33 | \$249,939 | [\$186,179 to \$320,234] |
| | | 50 | \$259,994 | [\$195,032 to \$343,611] |
| | | 67 | \$100,067 | [\$76,835 to \$125,803] |
| Vaginal | Female | 33 | \$269,164 | [\$195,013 to \$345,871] |
| | | 50 | \$215,222 | [\$162,357 to \$276,368] |
| | | 67 | \$56,614 | [\$43,953 to \$71,296] |
| Vulval | Female | 33 | \$260,501 | [\$190,626 to \$332,013] |
| | | 50 | \$190,397 | [\$145,529 to \$236,547] |
| | | 67 | \$48,825 | [\$38,310 to \$59,700] |

The benefit of vaccination, defined as the total avoided lost income less the cost of the vaccine, was highest for cervical cancer, with $1,347 [95% UI: $848 to $1,780] saved for each vaccinated girl. This rose to $1,397 [$895 to $1,838] when including the risk of income loss from other cancers. In boys, the avoided income losses did not outweigh the cost of vaccination, with an overall cost of $62 [$48 to $76]. These results are illustrated in Figure 5.

**Figure 5:**
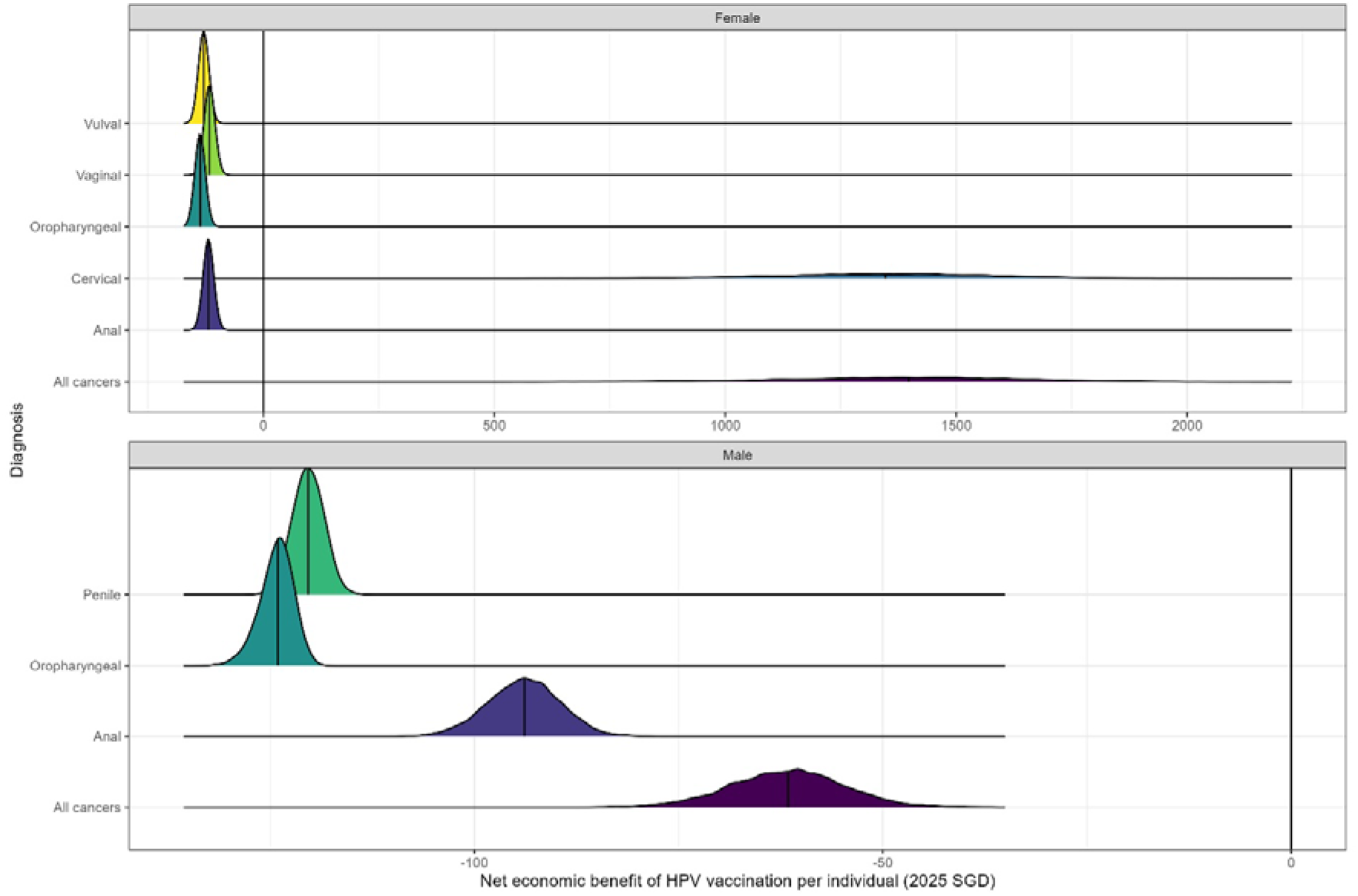
Probability-adjusted value of lost income when accounting for the net cost of each individual cancer, as well as all HPV-associated cancers together (bottom row), after subtracting vaccine costs.

### Sensitivity analysis results and extrapolation

As of 2024, there were 20,649 females and 21,432 males aged 15 in Singapore. Herd immunity would slightly increase the income protection benefits among vaccinated individuals, but would substantially increase the benefits among unvaccinated individuals. Assuming 80% vaccine coverage, the total cost of the policy per year would be $4.7m but returning a benefit of $29.1m [$18.9m to $38.3m]. This would yield a return on investment of $24.4m, or $5.2 gained per $1 spent.

## Discussion

Our results show that bivalent vaccination for HPV is a prudent strategy that parents and policymakers should strongly consider for economic purposes in both male and female school-aged children. While the prevalence of non-cervical cancers is relatively low in Singapore, the economic consequences of diagnosis at younger ages are severe, and the scale of mortality and unemployment burden following diagnosis can be a large proportion of overall lifetime income that can be mitigated by vaccination. While income losses alone are not sufficient for cost savings in boys, our model can be included in subsequent HPV cost-effectiveness research to estimate the broader economic costs of HPV-related cancers. Reducing bivalent vaccine costs to $76 would make vaccination cost-neutral in boys and increase savings in girls.

### Comparison with existing health-economic studies and policy considerations

There have been several studies of HPV vaccination cost-effectiveness in Singapore.^12–14^ The most comprehensive, by Wahab et al. (2023), showed that bivalent vaccination may be cost-effective in boys depending on the discount rate and vaccine cost.^16^ The authors noted that, while not cost-effective, reducing the total cost of the nonavalent vaccine over 2 doses to below $125 may be sufficient to qualify as cost-effective. Our study looked solely at the bivalent vaccine, but our results suggest that the overall cost of vaccines in a public healthcare system may be significantly reduced if considering the value of lost income. Future research must, however, consider the analytical perspective of such a model, as it has been argued that income losses are already accounted for by utility estimates.^37^

Our results align with findings from Europe, where Bencina et al (2024) found that cervical cancer was the greatest contributor to income losses, but oropharyngeal cancer had the greatest losses per diagnosis.^38^ This is likely because both their study and ours utilised many of the same model parameters. Hofmarcher et al (2020) also evaluated indirect costs of cancer in Europe, noting that indirect costs comprised around a third of the total economic burden of cancer in high-income countries including the United Kingdom, Germany and France.^39^ Assuming a roughly comparable split of direct and indirect costs, extrapolating these results to Singapore suggests that HPV vaccination would be net cost-saving for cancer in both boys and girls.

In addition to savings from avoided income losses, HPV vaccination could be an effective measure to reduce the cost of cancer surveillance. Chesson et al (2012) investigated the potential savings from avoidable cervical cancer screening costs due to preventing HPV-associated disease.^40^ Approximately 82% of the direct costs of HPV were related to cervical cancer screening; policymakers may wish to consider that cancer-related costs are only a fraction of the overall economic burden of HPV that can be addressed by vaccination.

The income losses due to oropharyngeal cancer diagnosis may be a relevant consideration for oral health policymakers. While parents may be aware of the relationship between HPV and cervical cancer, reluctance around dental administration of HPV vaccines may be tied to a lack of understanding of the role of HPV vaccination in oral health.^41^ This may be mitigated by framing vaccination in the context of oropharyngeal cancer prevention, as cancer prevention is generally considered the best reason for HPV vaccination by parents.^42^ Our results showed that the high burden of oropharyngeal cancer diagnosis may be a motivating explanation for parents to support vaccination by dental providers, especially if endorsed by governments and professional associations.

### Limitations

Our study solely assessed the costs to an individual in terms of lost lifetime income. This means that our study may be less relevant to healthcare decision-makers compared to traditional cost-effectiveness studies. However, our study demonstrates that even when individuals are insulated from direct healthcare costs, they are likely to bear an additional burden of losses in lifetime income, which may be severe over the life course.

We applied a similar approach to prior studies of HPV-associated cancer epidemiology in which HPV-related tumours of the primary site, rather than the pathological definition, were used to identify cases.^43^ The limitation is mitigated by the use of attributable fractions within the model, although they assume similar rates of HPV between our population and those in the published literature. Similarly, rates of cancer-related unemployment and sick leave may differ between Singapore and other populations. Sampling was used to incorporate our uncertainty of these model parameters.

Our analysis only tested the benefit of bivalent vaccination in Singapore, and applied average diagnosis and mortality rates from the past 30 years as input parameters for 2024 census estimates. These may limit study generalisability, and we encourage the secondary use of our data and code to inform subsequent work on this topic. In particular, our model was an economic, rather than epidemiological model, and was not able to capture population infection dynamics. Incorporating our results within epidemiological or microsimulation models may be able to form a more nuanced picture of income losses resulting from infection.

## Conclusion

Vaccination against HPV is an effective strategy that can protect against income losses, and parents should consider HPV vaccination for both health- and income-related reasons.

## Data Availability

All data produced by the study are available at https://github.com/robinblythe/HPV_CEA.

## Acknowledgements

The authors would like to thank the Singapore National Registry of Diseases Office for facilitating registry data access and ensuring the non-identifiability of extracted data.

